# Beneficial changes in the gut microbiome of patients with multiple sclerosis after consumption of Neu-REFIX B-glucan in a clinical trial

**DOI:** 10.1101/2023.09.07.23295172

**Authors:** Vidyasagar Devaprasad Dedeepiya, Chockanathan Vetrievel, Nobunao Ikewaki, Naoki Yamamoto, Hiroto Kawashima, Koji Ichiyama, Rajappa Senthilkumar, Senthilkumar Preethy, Samuel JK Abraham

## Abstract

**Background:** Multiple sclerosis (MS) is a debilitating demyelinating disease and recent evidences are giving cues towards correlation of disease severity to gut microbiome dysbiosis. However, there haven’t been any reported interventions that beneficially modifies the gut microbiome to yield a clinically discernible improvement. Having earlier reported the clinical effects of a biological response modifier beta-glucan (BRMG) produced by the N-163 strain of *Aureobasidum pullulans*, commercially available as Neu-REFIX, which decreased the biomarkers of inflammation and produced beneficial immune-modulation in twelve MS patients in 60 days, we evaluated their gut microbiome in the present study.

**Methods:** Twelve patients diagnosed with MS participated in the study. Each consumed 16 g gel of the NEU-REFIX beta-Glucan for 60 days. Whole genome metagenomic sequencing was performed on the fecal samples before and after Neu-REFIX intervention.

**Results:** Post-intervention analysis showed that *Actinobacteria* followed by *Bacteroides* was the major family. Abundance of beneficial genera such as *Bifidobacterium, Collinsela, Prevotella, Lactobacillus* and species such as *Prevotella copri (p-value=0.4), Bifidobacterium longum (p-value=0.2), Faecalibacterium prausnitzii (p-value=0.06), Siphoviridae (p-value=0.06)* increased while inflammation associated genera such as *Blautia (p-value=0.06)*, *Ruminococcus (p-value=0.007)* and *Dorea (p-value = 0.03)* decreased in abundance.

**Conclusion:** Restoration of gut eubiosis in terms of both increase in abundance of the good microbiome and suppression of the harmful ones which also correlate with earlier reported clinical improvement in MS patients makes this Neu-REFIX beta-glucan, a potential disease modifying therapy (DMT) requiring larger studies for validation in MS and other auto-immune-inflammatory conditions where a safe intervention for immune modulation is vital.

## Introduction

Multiple sclerosis (MS) is a chronic autoimmune condition that causes demyelination, axonal loss, and inflammation of the central nervous system (CNS). There are 2.5 million people with MS worldwide and it imposes severe burden on the affected patients, their families and the society as a whole. Although the exact cause of MS is unknown, host genetics and environmental variables have been theorised to play a role. MS is a diverse illness that is influenced by both environmental and genetic factors, such as the relationship with HLA-DRB1*15:01 and vitamin D status, obesity, smoking, and Epstein-Barr virus (EBV) infection. MS patients’ dysregulated immune responses and aberrant metabolism raise the possibility that the disease’s pathogenesis involves a number of systems [1]. MS pathogenesis is influenced by changes in the peripheral immune system, blood brain barrier permeability, and intrinsic CNS immune cells (such microglia). Neurodegeneration and acute and chronic inflammation both occur over the course of the illness, with acute inflammation being more prominent during the relapsing phase. Although the question of whether MS is neurological or autoimmune by origin is still being debated, majority of genetic profiling demonstrates a preponderance of immune-mediated molecules that are activated during the illness process. Both innate and adaptive immune responses play a role, as shown by animal models and clinical findings in MS patients. Myeloid-derived macrophages and microglia are innate immune cells that play important roles in MS. Autoimmune CD4+ T cells, in particular Th1 cells that are reactive to myelin proteins, and CD8+ cytotoxic T cells are adaptive immune cells that are involved in MS. B cells have a significant role in the pathogenesis of human MS through the generation of proinflammatory cytokines and chemokines, antibody creation, and antigen presentation to T cells [1]. Multiple pathogenic mechanisms, including activated microglia, leptomeningeal inflammatory infiltrates causing subpial demyelination, mitochondrial dysfunction and oxidative injury driven by macrophages and microglia, are thought to contribute to the development of progressive MS [2].

In this background, the role of the gut microbiome which has a bi-directional influence on the central nervous system, immune and inflammation mediated processes is becoming increasingly associated with the initiation and progression of MS. Changes in the gut microbiota trigger immunological and physiological changes in the host, which are crucial for preserving the gut microbiota’s homeostasis. The intricacy of the gut environment, as currently understood, points to the importance of dysbiosis, which is defined as changes in the microbiome’s composition acting as a potential disease-causing mechanism. Dysbiosis can impact immune reactions to the microbiota, but it can also negatively impact the integrity of the epithelia, which form cellular barriers necessary to preserve the health of the intestine and the central nervous system (CNS) [3].

We have earlier reported the beneficial effects of N-163 strain of *Aureobasidium pullulans*, produced biological response modifier beta-glucan (BRMGs), trade name - Neu-REFIX on the gut microbiome in pre-clinical and clinical studies in Non-alcoholic steatohepatitis (NASH) [4] and Duchenne muscular dystrophy (DMD) [5]. These effects included the regulation of gut dysbiosis, enhancement of butyrate producers, and metabolites, including endogenous butyrate production and amino acids. Importantly, these results correlated with the improvement in the clinical parameters of relevance [6,7].

The clinical improvement in immunological and inflammatory parameters such as decrease in IL-6, improvement in CD4+ve, CD19+ve, CD3+ve, and CD8+ ve cell count, increase in Lymphocyte to C-reactive protein ratio (LCR), Leukocyte to CRP ratio (LeCR) and a decrease in Neutrophil to Lymphocyte ratio (NLR) apart from improvement in Kurtzke’s expanded disability severity score (EDSS) have been studied in the clinical trial done in MS patients [8].

In the same patients, in the current study, we have evaluated the effects of this Neu-REFIX on their gut microbiome.

## Methods

This trial was an open label, prospective, non-randomised, non-comparative single arm clinical study. The study was of 60 days duration. The inclusion and exclusion criteria are available in the study on clinical parameters reported earlier [8]. Patients with MS diagnosed as per McDonald diagnostic criteria (2005) were included in the study.

Prior to the study, the patients’ treatment plans varied in terms of the medications they took and how long they were given to work, but the majority of them included steroids, muscle relaxants, and in the case of some patients, interferon beta-1b (IFNB) and immunomodulators like alemtuzumab, fingolimod, or natalizumab.

In addition to their above-mentioned standard treatment plan, the patients took two sachets of the N-163 strain of A.pullulans (each sachet of 8g gel contains 48 mg of active substance; trade name: Neu-REFIX) produced beta-glucan, every day along with a meal for 60 days.

Using a sterile faeces collection kit (OMNIgene Gut collection kit), faeces samples were taken from all the subjects at baseline and 60 days after the intervention. DNA extraction was performed and samples were stored at minus 80 °C until they were needed for analysis. Following the manufacturer’s instructions, total microbial DNA was extracted from each sample of faeces using the QIAamp DNA Mini Kit (Qiagen). An extraction control (negative buffer control) was used to extract each group of samples.

The samples were sequenced using Novaseq V1.5 with a read length of 151 bp. The samples were subjected to whole genome metagenome analysis. Initially, the reads were filtered for human DNA contamination. The alignment to human genome was around 0.01% - 1.6%. The filtered reads were further used downstream analysis. De novo metagenome assembly was carried out using Megahit assembler. Assembly was performed using de-Bruijn graph method.

The assembled scaffolds were taken for further downstream analysis. Open reading frame (ORF) prediction was done using Prodigal (v2.6.3). The assembled scaffolds from the de novo assembly was taken for the ORF prediction. The obtained ORFs were filtered using in house Perl scripts. ORFs of length below 200 bp were filtered out. The abundances at the phylum, genus and species level were evaluated. Pair-wise comparison phylum to species taxapair-wise comparison phylum to species taxa was performed

Statistical data were analyzed using GraphPad Prism software. The statistical significance level was set at 5%. Non-parametric tests such as Mann Whitney test for independent measures were employed were used for metagenomic sequencing data, including taxonomy.

## Results

Gut bacterial gene richness were similar pre-and post-intervention. Post-intervention, the abundance of *Actinobacteria*, *Bacteroidetes* and *Proteobacteria* increased at genus level (Figure 1). At phylum level, post-intervention, *Bifidobacterium*, *Prevotella*, *Collinsella i*ncreased while *Blautia* and Dorea decreased (Figure 1). When individual species and genera were analysed (Figure 2), there was increase in mean abundance of *Bifidobacterium longum* from 1.01 ± 0.8 to 1.3 ± 0.8 (p-value= 0.2), *Prevotella copri* from 6.529 ± 11.14 to 7.22 ± 12.13 (p-value= 0.4), *Faecalibacterium prausnitzii* from 4.7 ± 4.2 to 7.29 to ±4.54 (p-value= 0.06)*, Siphoviridae sp* from 2.79 ± 1.8 to 2.97 ± 1.9 (p-value= 0.06) (Figure 3). The mean abundance of *Blautia* decreased from 0.24 ± 0.l8 to 0.01 ± 0.05 (p-value = 0.003), *Dorea* decreased from 0.63 ± 0.77 to 0.33 ± 0.73 (p-value = 0.03) and *Ruminococcus* decreased from 0.93 ± 0.56 to 0.37 ± 0.31 (p-value = 0.007) post intervention (Figure 4).

**Figure 1:**
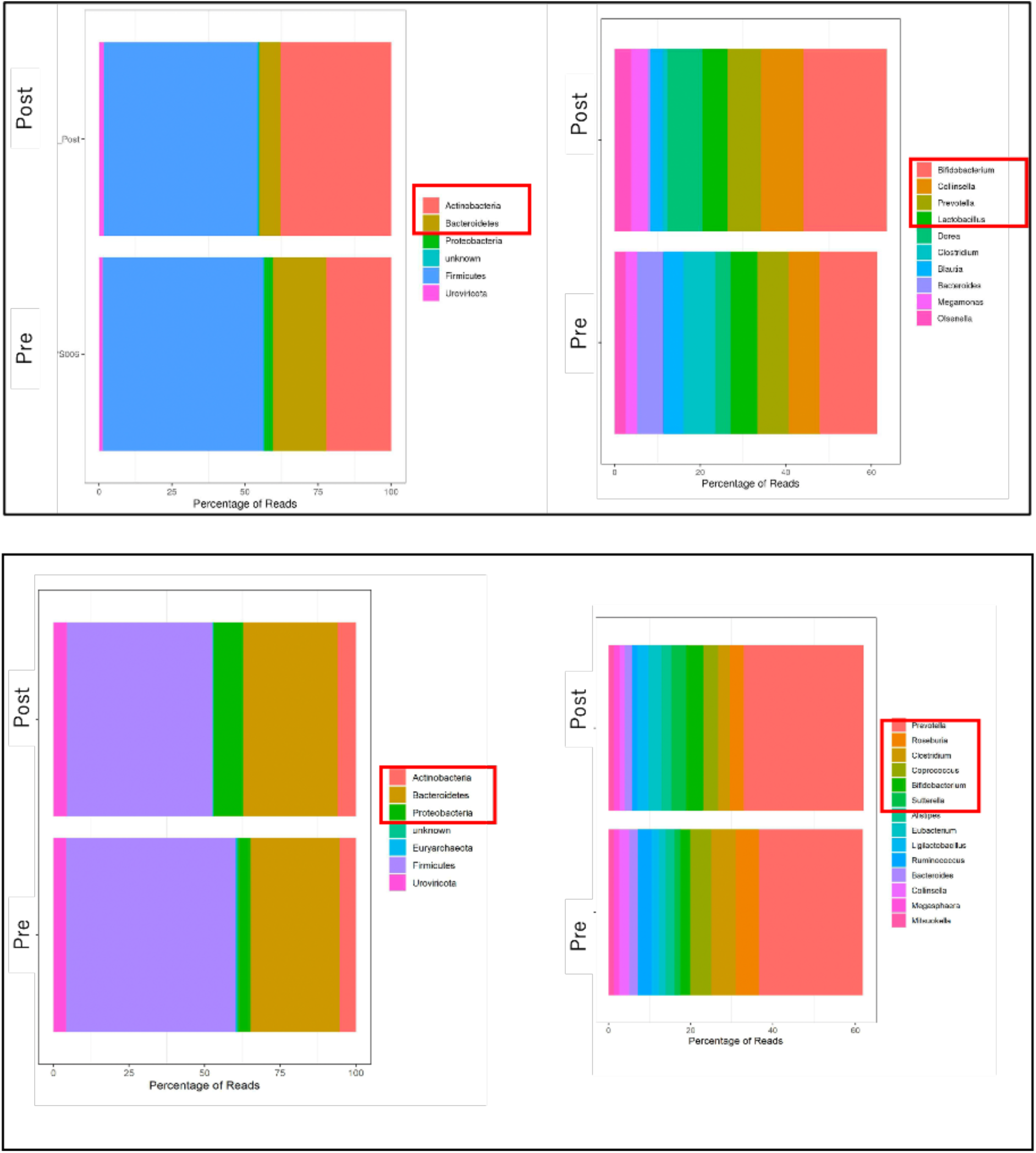
Relative abundance at phylum and genus level.

**Figure 2:**
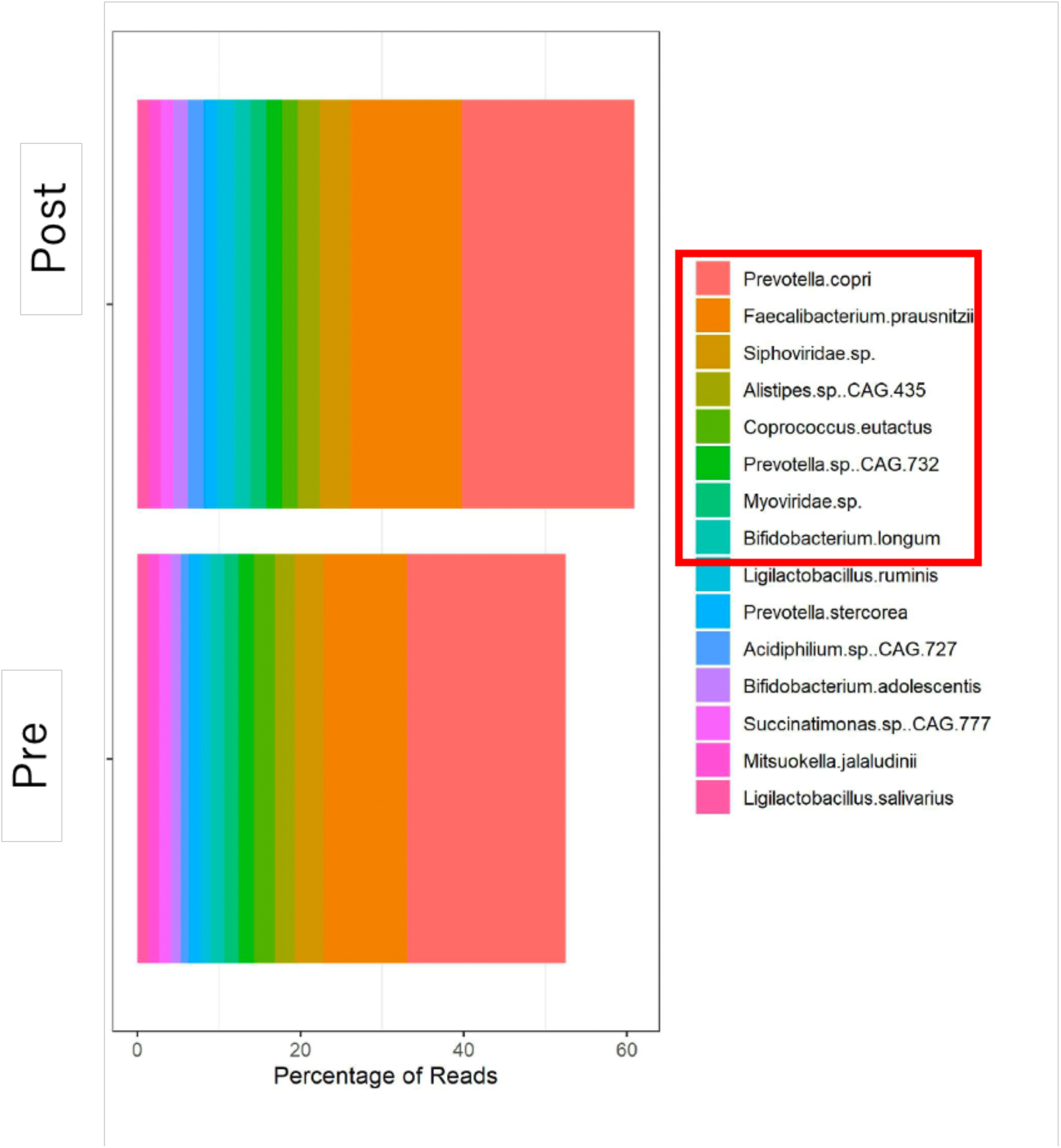
Relative abundance at species level.

**Figure 3:**
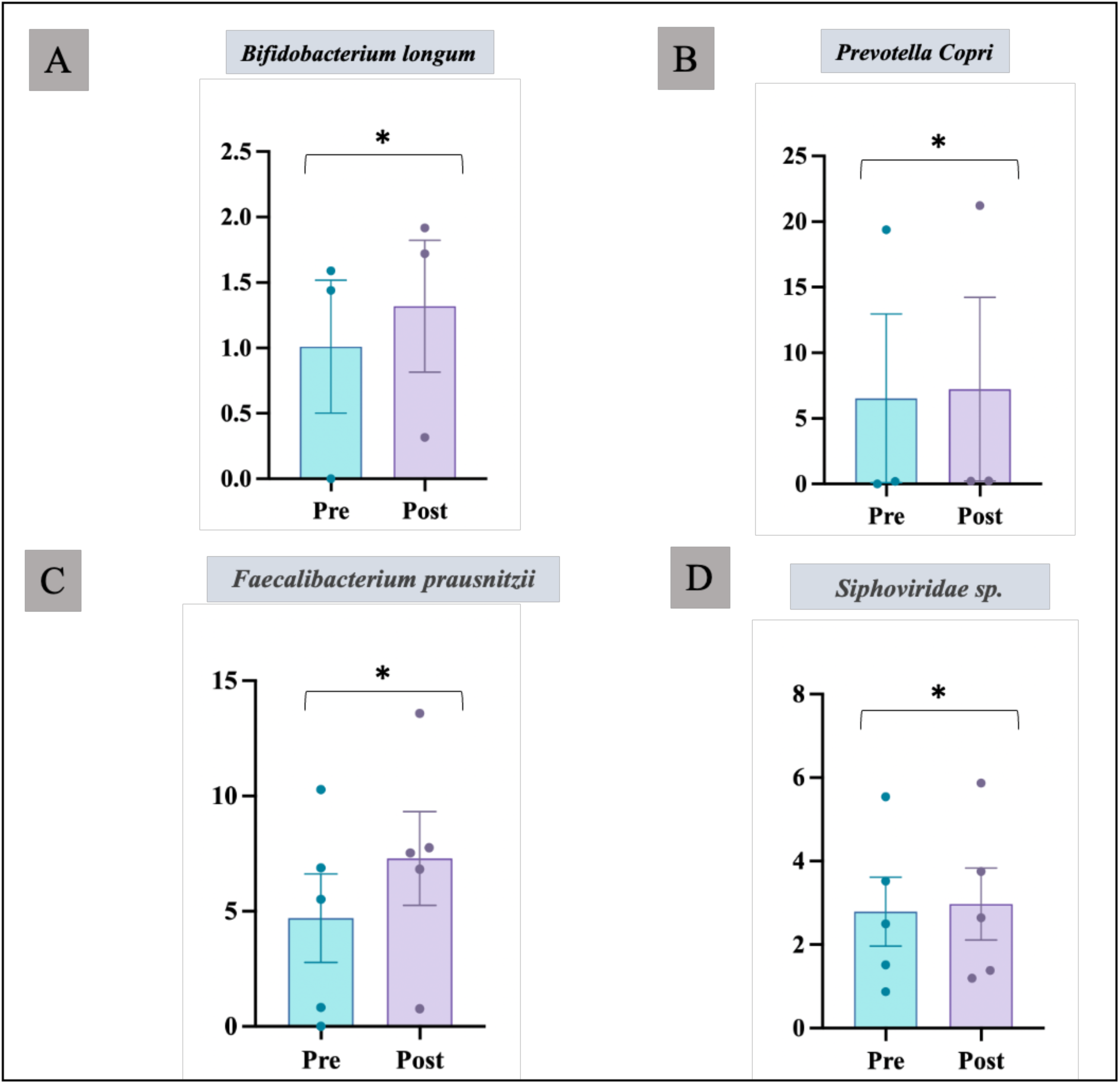
Increase in abundance of beneficial microorganisms: A. *Bifidobacterium longum*, B. *Prevotella Copri, C. Faecalibacterium prausnitzii and D. Siphoviiridae sp*. *p-value > 0.05 (n.s)

**Figure 4:**
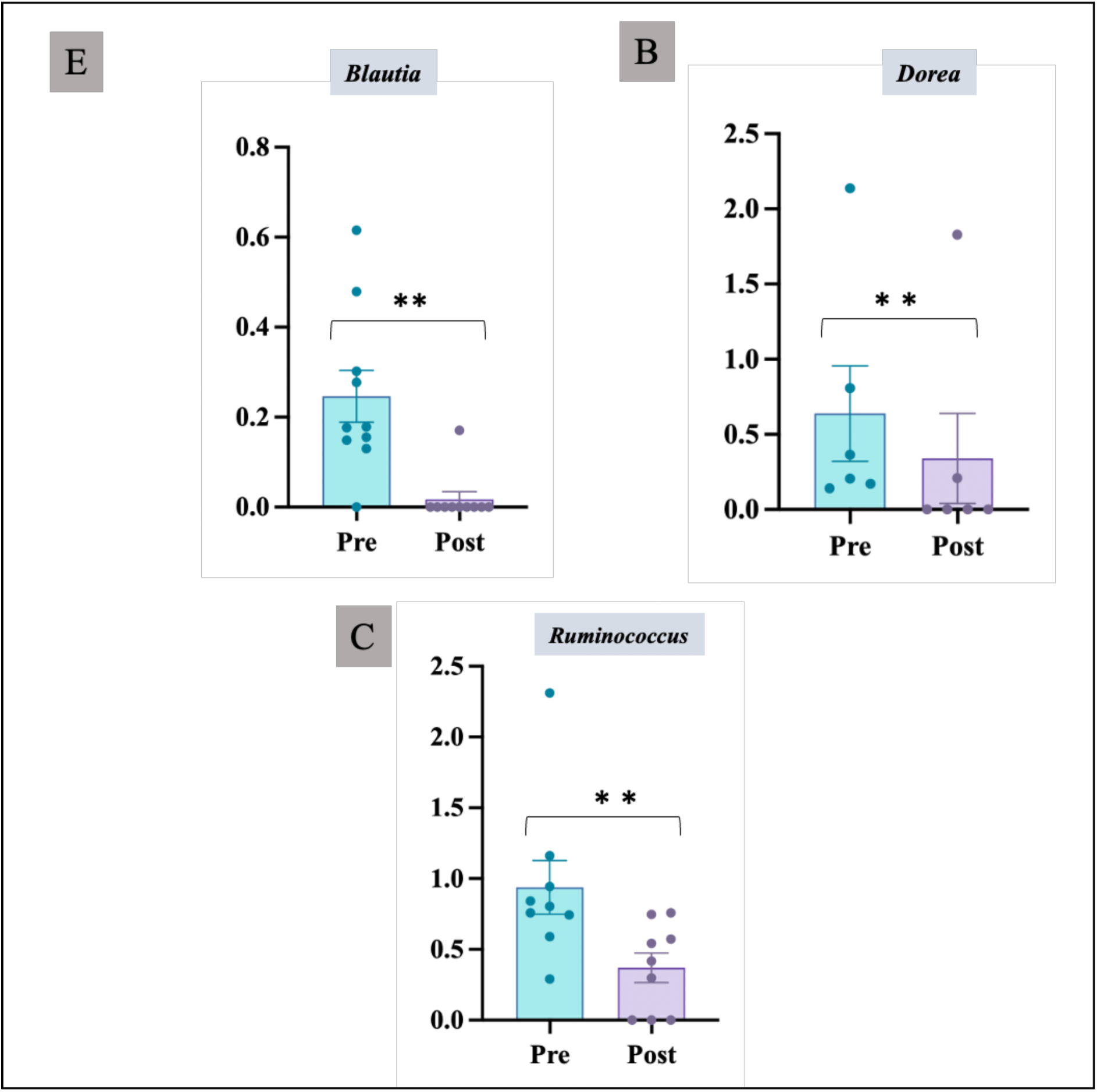
Decrease in abundance of harmful microorganisms: A. *Blautia*, C. *Dorea* and D. *Ruminococcus*, post NEU-REFIX Beta-glucan intervention **p-value < 0.05 (significant)

## Discussion

Multiple sclerosis (MS) is one of the rare demyelinating, devastating diseases which has very complex pathogenesis, ambiguous aetiology and extensive heterogeneity in terms of clinical features, genetics, pathogenesis and responsiveness to treatments. The clinical course of MS is either relapsing-remitting (RR), which represents around 60% of prevalent cases, or progressive [9]. About 10% of MS cases have a primary progressive (PP) course, while transition to the secondary progressive (SP) phase occurs in around half of RR MS. Standard care of treatments include DMTs such as interferon beta (IFNB) 1-a and 1-b, glatiramer acetate (GA), mitoxantrone, natalizumab, fingolimod, teriflunomide, dimethyl fumarate, and alemtuzumab, off label use of azathioprine, cyclophosphamide, methotrexate and rituximab but most of these treatments are associated with increased risk of serious adverse events, which may even be fatal in rare cases [9]. Therefore, there is a need to develop safe DMTs. With reference to the aetiology of the disease, there are several hypothesis of environmental, genetic factors, such as the relationship with HLA-DRB1*15:01 and vitamin D status, obesity, smoking, auto-immune and Epstein-Barr virus (EBV) infection [1]. However, the DMTs mentioned above are not specific and efficacy still is largely limited with neurological disability continuing to worsen over time. Specific targeted immune-suppression is also not possible with these DMTs.

In this background, the only solace is the correlation of gut microbiome and disease severity which opens a new avenue to explore gut microbiome based approaches as the pathophysiology of MS appears to be significantly influenced by the gut flora, and it appears to be involved in controlling the host’s immune system, affecting the BBB’s integrity and functionality, causing autoimmune demyelination, directly engaging many CNS cell types. In earlier studies, when compared to healthy individuals and MS patients in the remittent phase, people with MS in the relapse phase exhibited increase in *firmicutes* and a decline in the phylum *Bacteroidetes* [10–12]. Additionally, it was discovered that *Prevotella* had decreased. *Prevotella* has been shown to reduce the anti-inflammatory metabolite propionate and therefore its decrease in MS is harmful. Genera like *Blautia, Ruminococcus* and *Dorea* have been reported to be in greater abundance in patients with MS compared to healthy controls [13]. Instead, the prevalence of *Bacteroidetes* genera like *Parabacteroides*, *Bacteroides*, and *Prevotella* was reduced [10–12]. Because they may activate IFN, metabolise sialic acids, and breakdown mucin, certain species of *Dorea* may be pro-inflammatory, according to research by Schirmer et al [13]. In light of this, *Dorea* could serve as an illustration of a bacterium that shows pro or anti-inflammatory functions depending on the nearby gut flora and/or nutrients that are accessible. In fact, since *Blautia* uses the gases generated by *Dorea*, a higher concentration of *Dorea* in MS patients may encourage the formation of *Blautia* [12]. *Bifidobacteria* may play a protective function in MS, according to a number of lines of evidence. Numerous strategies, such as a change in lipid profile, stimulation of Treg differentiation, and modification of the Th1/Th2 balance, might be the mechanisms underlying such a protective impact [14]. Due to its function in butyrate generation, *Faecalibacterium* is linked to a decreased inflammatory state, and methods to increase *Faecalibacterium* abundance have been proposed as potential treatments for ulcerative colitis patients. Earlier reports have shown that subjects with increased *Caudovirales* and *Siphoviridae* levels in the gut microbiome had better performance in executive processes and verbal memory [15]. All these studies have only reported a correlation of gut microbiome to the disease severity of the disease. However to our knowledge, till date, there has been no study which has reported an intervention that has relevance to clinical outcome and the gut microbiome. Herein comes our study’s results wherein a clinically encouraging outcome in terms of EDSS, inflammatory and immune-modulatory biomarkers which was earlier reported [8], which in follow-up of evaluation of gut microbiome in our present study, a striking increase in beneficial bacteria with a significant reduction in harmful bacteria, we have been able to prove. In our study there has been an increase in phylum *Bacteroidetes* and *Prevotella* post-intervention. There is a reversal effect of decrease in genera like *Blautia, Ruminococcus* and *Dorea* with increase in *Bacteroidetes, Bifidobacterium*, *Faecalibacterium* and *Siphoviridae* levels in our study which is beneficial.

While the relevance of gut microbiome has been reported in several earlier studies, manipulating it beneficially involves methods like fecal microbiota transplantation (FMT) or probiotics which essentially are exogenous supplementation of beneficial microbiome [16]. However, they don’t essentially address the harmful bacterial colonies and their metabolites and the environment. Our approach does both; decreasing the abundance of harmful organisms while increasing the beneficial ones more in an endogenous manner apart from immune modulation and producing clinically relevant outcome. Further, the Neu-REFIX Beta-Glucan is safe and is easily consumable making it a potential disease modifying adjunct for MS both clinically and in relevance to the gut microbiome.

Limitations of the study include small sample size, heterogenous presentation of MS, geographical influences as this study consisted of participants of Indian ethnicity alone while western populations have reported a higher prevalence of MS. Therefore further validation in different populations with global multi-centric clinical trials is needed. Now having found a small step in beneficial reconstituting the gut microbiome in this pilot clinical study, an in depth exploration is needed to find if gut dysbiosis is a cause or effect in the background of underlying inflammation in response to an unknown antigen. This question may have been partially answered in this study because Neu-REFIX beta-glucan has shown beneficial effects of restoring the gut microbiome and also the improvement in clinical parameters could be attributed as an effect. Going by the earlier reports of gut dysbiosis caused by specific drugs in neurological illness [17,18], in line with those evidences, whether similar drugs are part of MS regimen have to be looked into or diseases that have direct correlation to gut dysbiosis when subjected to a clinical study, the appropriate drug or molecule should be studied for implications on gut microbiome as well.

## Conclusion

In this pilot clinical study of patients with MS, who have reported improvement in clinical biomarkers of inflammation and immune-modulation such as decrease in IL-6, improvement in CD4+ve, CD19+ve, CD3+ve, and CD8+ ve cell count, increase in Lymphocyte to C-reactive protein ratio (LCR), Leukocyte to CRP ratio (LeCR) and a decrease in Neutrophil to Lymphocyte ratio (NLR) apart from improvement in EDSS [8], their gut microbiome after intervention has shown a significant correlation. Such correlation is very well falling in line with the reported indicators of severity of clinical findings, wherein specific gut microbiome associated with relapse of disease, have decreased after Neu-REFIX beta-glucan in the present study. This is one of its kind report in the literature yielding a dual advantage improving the good microbiome and suppressing the harmful ones, making us recommend Neu-REFIX beta-glucan a potential DMT requiring larger studies for validation not only in MS but also other auto-immune-inflammatory conditions.

## Data Availability

All data generated or analysed during this study are included in the article itself.

## Acknowledgements

The authors thank

1. The Government of Japan and the Prefectural Government of Yamanashi for a special loan and M/s Yamanashi Chuo Bank for processing the transactions.
2. Ms. Ann Gonsalvez and members of Multiple Sclerosis Society of India (MSSI)- Chennai Chapter for their coordination with the participants of the study.
3. Dr. Malcolm Jeyaraj, Neurologist for his clinical inputs and evaluation of the patients.
4. Prof. Dr. R. Raveendran, Dept. of Pharmacology, Jawaharlal Institute of Postgraduate Medical Education & Research (JIPMER), Pondicherry, India for providing his valuable inputs on the statistical analysis of the study.
5. Dr. Ragaroobine, Mr. Rajmohan from Nichi-In Centre for Regenerative Medicine (NCRM) for their assistance with data collection.
6. Mr. Masato Onaka, Mr. Yasushi Onaka of Sophy Inc, Kochi, Japan for technical advice.
7. Ms. Yoshiko Amikura and staff of GN Corporation Co Ltd, Japan for their liaison assistance with the conduct of the study.

## References

1. Mansilla MJ, Presas-Rodríguez S, Teniente-Serra A, González-Larreategui I, Quirant-Sánchez B, Fondelli F, Djedovic N, Iwaszkiewicz-Grześ D, Chwojnicki K, Miljković Đ, Trzonkowski P, Ramo-Tello C, Martínez-Cáceres EM. Paving the way towards an effective treatment for multiple sclerosis: advances in cell therapy. Cell Mol Immunol. 2021 Jun;18(6):1353–1374.

2. Lassmann H. Multiple Sclerosis Pathology. Cold Spring Harb Perspect Med. 2018 Mar 1;8(3):a028936.

3. Freedman SN, Shahi SK, Mangalam AK. The “Gut Feeling”: Breaking Down the Role of Gut Microbiome in Multiple Sclerosis. Neurotherapeutics. 2018 Jan;15(1):109–125.

4. Preethy S, Ikewaki N, Levy GA, Raghavan K, Dedeepiya VD, Yamamoto N, Srinivasan S, Ranganathan N, Iwasaki M, Senthilkumar R, Abraham SJK. Two unique biological response-modifier glucans beneficially regulating gut microbiota and faecal metabolome in a non-alcoholic steatohepatitis animal model, with potential applications in human health and disease. BMJ Open Gastroenterol. 2022 Sep;9(1):e000985.

5. Raghavan K, Dedeepiya VD, Yamamoto N, Ikewaki N, Iwasaki M, Dinassing A, Senthilkumar R, Preethy S, Abraham SJK. Randomised trial of Aureobasidium pullulans-produced beta 1,3-1,6-glucans in patients with Duchenne muscular dystrophy: Favourable changes in gut microbiota and clinical outcomes indicating their potential in epigenetic manipulation. medRxiv 2022.12.09.22283273; doi: 10.1101/2022.12.09.22283273

6. Ikewaki N, Levy GA, Kurosawa G, Iwasaki M, Dedeepiya VD, Vaddi S, Senthilkumar R, Preethy S, Abraham SJK. Hepatoprotective Effects of Aureobasidium pullulans Derived β 1,3-1,6 Glucans in a Murine Model of Non-alcoholic Steatohepatitis. J Clin Exp Hepatol. 2022 Nov-Dec;12(6):1428-1437.

7. Raghavan K, Dedeepiya VD, Srinivasan S, Pushkala S, Bharatidasan SS, Ikewaki N, Iwasaki M, Senthilkumar R, Preethy S, Abraham SJK. Beneficial immune-modulatory effects of the N-163 strain of Aureobasidium pullulans-produced 1,3-1,6 Beta glucans in Duchenne muscular dystrophy: Results of an open-label, prospective, exploratory case-control clinical study. IBRO Neurosci Rep. 2023;15: 90-99.

8. Dedeepiya VD, Vetrievel C, Ikewaki N, Ichiyama K, Yamamoto N, Kawashima H, Bharatidasan SS, Srinivasan S, Senthilkumar R, Preethy S, Abraham S. Improvement in Expanded Disability Status Scale (EDSS) and anti-inflammatory parameters in patients with multiple sclerosis following oral consumption of N-163 strain of Aureobasidium pullulans produced beta glucan in a pilot clinical study. medRxiv 2023.05.14.23289953v2; doi: 10.1101/2023.05.14.23289953

9. Gajofatto A, Benedetti MD. Treatment strategies for multiple sclerosis: When to start, when to change, when to stop? World J Clin Cases. 2015 Jul 16;3(7):545–55

10. Schepici G, Silvestro S, Bramanti P, Mazzon E. The Gut Microbiota in Multiple Sclerosis: An Overview of Clinical Trials. Cell Transplant. 2019 Dec;28(12):1507–1527.

11. Chen J, Chia N, Kalari KR, Yao JZ, Novotna M, Soldan MMP, Luckey DH, Marietta EV, Jeraldo PR, Chen XF, Weinshenker BG, et al. Multiple sclerosis patients have a distinct gut microbiota compared to healthy controls. Sci Rep Uk. 2016;6:28484.

12. Shahi SK, Freedman SN, Mangalam AK. Gut microbiome in multiple sclerosis: The players involved and the roles they play. Gut Microbes. 2017 Nov 2;8(6):607–615.

13. Schirmer M, Smeekens SP, Vlamakis H, Jaeger M, Oosting M, Franzosa EA, Horst RT, Jansen T, Jacobs L, Bonder MJ, Kurilshikov A, Fu J, Joosten LAB, Zhernakova A, Huttenhower C, Wijmenga C, Netea MG, Xavier RJ. Linking the Human Gut Microbiome to Inflammatory Cytokine Production Capacity. Cell. 2016 Dec 15;167(7):1897.

14. Cristofori F, Dargenio VN, Dargenio C, Miniello VL, Barone M, Francavilla R. Anti-Inflammatory and Immunomodulatory Effects of Probiotics in Gut Inflammation: A Door to the Body. Front Immunol. 2021 Feb 26;12:578386.

15. Mayneris-Perxachs J, Castells-Nobau A, Arnoriaga-Rodríguez M, Garre-Olmo J, Puig J, Ramos R, Martínez-Hernández F, Burokas A, Coll C, Moreno-Navarrete JM, Zapata-Tona C, Pedraza S, Pérez-Brocal V, Ramió-Torrentà L, Ricart W, Moya A, Martínez-García M, Maldonado R, Fernández-Real JM. Caudovirales bacteriophages are associated with improved executive function and memory in flies, mice, and humans. Cell Host Microbe. 2022 Mar 9;30(3):340–356.e8.

16. Borody T, Leis S, Campbell J, Torres M, Nowak A. Fecal microbiota transplantation (FMT) in multiple sclerosis (MS). Am J Gastroenterol. 2011;106:S352–S352.

17. Ternák G, Kuti D, Kovács KJ. Dysbiosis in Parkinson’s disease might be triggered by certain antibiotics. Med Hypotheses. 2020 Apr;137:109564.

18. Ternák G, Németh M, Rozanovic M, Márovics G, Bogár L. Antibiotic Consumption Patterns in European Countries Are Associated with the Prevalence of Parkinson’s Disease; the Possible Augmenting Role of the Narrow-Spectrum Penicillin. Antibiotics (Basel). 2022 Aug 23;11(9):1145.

